# Randomized Controlled Trial of Early Outpatient COVID-19 Treatment with High-Titer Convalescent Plasma

**DOI:** 10.1101/2021.12.10.21267485

**Authors:** David J. Sullivan, Kelly A. Gebo, Shmuel Shoham, Evan M. Bloch, Bryan Lau, Aarthi G. Shenoy, Giselle S. Mosnaim, Thomas J. Gniadek, Yuriko Fukuta, Bela Patel, Sonya L. Heath, Adam C. Levine, Barry R. Meisenberg, Emily S. Spivak, Shweta Anjan, Moises A. Huaman, Janis E. Blair, Judith S. Currier, James H. Paxton, Jonathan M. Gerber, Joann R. Petrini, Patrick B. Broderick, William Rausch, Marie Elena Cordisco, Jean Hammel, Benjamin Greenblatt, Valerie C. Cluzet, Daniel Cruser, Kevin Oei, Matthew Abinante, Laura L. Hammitt, Catherine G. Sutcliffe, Donald N. Forthal, Martin S. Zand, Edward R. Cachay, Jay S. Raval, Seble G. Kassaye, E. Colin Foster, Michael Roth, Christi E. Marshall, Anusha Yarava, Karen Lane, Nichol A. McBee, Amy L. Gawad, Nicky Karlen, Atika Singh, Daniel E. Ford, Douglas A. Jabs, Lawrence J. Appel, David M. Shade, Stephan Ehrhardt, Sheriza N. Baksh, Oliver Laeyendecker, Andrew Pekosz, Sabra L. Klein, Arturo Casadevall, Aaron A.R. Tobian, Daniel F. Hanley

## Abstract

**BACKGROUND:** The efficacy of polyclonal high titer convalescent plasma to prevent serious complications of COVID-19 in outpatients with recent onset of illness is uncertain.

**METHODS:** This multicenter, double-blind randomized controlled trial compared the efficacy and safety of SARS-CoV-2 high titer convalescent plasma to placebo control plasma in symptomatic adults ≥18 years positive for SARS-CoV-2 regardless of risk factors for disease progression or vaccine status. Participants with symptom onset within 8 days were enrolled, then transfused within the subsequent day. The measured primary outcome was COVID-19-related hospitalization within 28 days of plasma transfusion. The enrollment period was June 3, 2020 to October 1, 2021.

**RESULTS:** A total of 1225 participants were randomized and 1181 transfused. In the pre-specified modified intention-to-treat analysis that excluded those not transfused, the primary endpoint occurred in 37 of 589 (6.3%) who received placebo control plasma and in 17 of 592 (2.9%) participants who received convalescent plasma (relative risk, 0.46; one-sided 95% upper bound confidence interval 0.733; P=0.004) corresponding to a 54% risk reduction. Examination with a model adjusting for covariates related to the outcome did not change the conclusions.

**CONCLUSION:** Early administration of high titer SARS-CoV-2 convalescent plasma reduced outpatient hospitalizations by more than 50%. High titer convalescent plasma is an effective early outpatient COVID-19 treatment with the advantages of low cost, wide availability, and rapid resilience to variant emergence from viral genetic drift in the face of a changing pandemic.

**Trial Registration:** ClinicalTrials.gov number, NCT04373460.

## INTRODUCTION

Severe Acute Respiratory Syndrome coronavirus 2 (SARS-CoV-2) which causes coronavirus disease 2019 (COVID-19) has an eight percent United States hospitalization rate, and over 5 million deaths worldwide. To date most therapies have targeted disease progression or death in hospitalized patients. Three outpatient monoclonal antibody (mAb) therapies received Food and Drug Administration (FDA) Emergency Use Authorization (EUA), after data showed reductions in disease progression and hospitalizations when given within 5 to 7 days of illness onset^1-3^. Alternative outpatient therapies are needed, particularly in settings where mAb therapy is either unavailable (e.g. in low and middle income countries)^4^, scarce, or ineffective (i.e. in the context of mAb-resistant variants)^5^.

COVID-19 convalescent plasma (CCP) is safe in hospitalized populations^6,7^. High titer CCP given early in the hospital also reduced deaths by 50%^8^ but the limited evidence from randomized clinical trials has been mixed with some studies showing efficacy in reducing mortality^9,10^ and others not ^11-14^. Generally, CCP is most effective when provided early and high-titer^15^. However, outpatient randomized trial data are limited^16^. Outpatient CCP use showed 48% efficacy when used within 3 days of mild COVID-19 symptoms in a trial conducted in Argentina^17^, whereas a study of CCP among emergency department (ED) patients at high risk for progression of COVID-19 was halted due to perceived futility^18^.

We sought to determine if high titer CCP (greater than 1:320 SARS-CoV-2 spike protein titers), transfused within 9 days of symptom onset would be effective at preventing hospitalization in adults over age 18 regardless of comorbidities and COVID-19 vaccine receipt.

## METHODS

### TRIAL DESIGN AND OVERSIGHT

The Convalescent Plasma to Limit SARS-CoV-2 Associated Complications (CSSC-004) Study was a double-blind randomized controlled trial comparing high titer CCP to placebo control plasma. The study was conducted under an FDA Investigational New Drug application sponsored by Johns Hopkins University (IND 19725). Enrollment sites and investigators are listed in supplementary appendix.

Johns Hopkins served as the single-IRB (sIRB). For the Center for American Indian Health sites, the protocol was also independently reviewed and approved by the Navajo Nation Health Human Research Review Board and the National Indian Health Service IRB. The protocol was also approved by the Department of Defense (DoD) Human Research Protection Office (HRPO). An independent medical monitor reviewed all Serious Adverse Events (SAE) and an independent, masked, three physician, panel adjudicated COVID-19 related hospitalizations and severity scores (Appendix). An independent Data Safety Monitoring Board (DSMB) provided interim safety and efficacy reviews (Appendix). The trial was conducted in accordance with the principles of the Declaration of Helsinki, International Council for Harmonization Good Clinical Practice guidelines, and all applicable regulatory requirements.

The authors responsible for trial design, data assembly, analysis and manuscript writing are listed in the Appendix. All authors vouch for trial protocol adherence, the completeness and accuracy of the data and analyses.

### PARTICIPANTS

We enrolled SARS-CoV-2 positive participants age ≥18 years within 8 days of symptom onset and transfusion by day 9. Exclusion criteria included prior COVID-19 hospitalization or planned hospitalization within 24 hours of enrollment, prior transfusion reactions, inability to comply with the protocol transfusion or follow up, or mAb receipt before enrollment. Persons who received a SARS-CoV-2 vaccine before or during follow-up, received corticosteroid treatment, or were pregnant or breast-feeding were eligible. All participants provided written informed consent.

### RANDOMIZATION AND INTERVENTION

After screening and consent, participants from all sites were randomized by a central web-based system using blocked assignments to receive ABO compatible control plasma or CCP (both∼250 mL) in a 1:1 ratio. Both investigational products were compatibility matched and then over labeled as “Thawed Plasma [volume] Store at 1-6°C; New Drug Limited by Federal (or United States) law to investigational use” thereby preserving verification codes. Control plasma or CCP was transfused over approximately one hour within 24 hours of enrollment followed by observation for 30 minutes.

Eligible donors were qualified with SARS-CoV-2 antibody (Euroimmun) with minimum titers of ≥1:320 as determined using a validated ELISA assay in a CLIA certified laboratory. The March 9, 2021 FDA EUA^19^ for high titer hospital CCP was used as the basis to further standardize units for study transfusion after July 2021. After qualification, the donor CCP was characterized for antibody levels by Euroimmun ratio at 1:101 and endpoint titers^20^.

### OUTCOMES

The measured primary outcome was cumulative COVID-19 related hospitalization incidence in CCP treatment versus control group by day 28. While death before hospitalization was part of a composite primary outcome, it did not occur in the study. Hence, the primary outcome is equivalent to COVID-19 hospitalization. COVID-19 relatedness for hospitalizations was adjudicated by a three physician panel masked to treatment group. Other pre-specified endpoints included hospital disease severity measured by ICU admission, invasive mechanical ventilation, or death in hospital.

### SAFETY ASSESSMENTS

Adverse events (AEs) were monitored throughout the study. Safety outcomes included transfusion-related SAEs manifested as severe transfusion reactions, acute respiratory distress syndrome and grade 3 or 4 adverse events. An independent medical monitor evaluated AEs, SAEs and changes in baseline safety laboratory values.

### STATISTICAL ANALYSIS

The statistical analysis plan was finalized before database lock and unmasking (Appendix). A sample size of 1280 was determined to have 80% power using a one-sided test to detect at least a 25% reduction in hospital risk and was inflated to 1344 to allow for potential loss to follow-up. We calculated the risk difference (RD) and the restricted mean survival time (expected mean time to hospitalization or death by 28 days) in a modified intention to treat (mITT) analysis, that excluded those who did not receive transfusion of study plasma. We estimated the cumulative incidence using the doubly robust estimator based upon targeted minimum loss-based estimator (TMLE)^21^. Analyses were adjusted for baseline variables potentially related to the outcome in order to increase estimate precision and account for potential dependent censoring^21^. To determine which pre-specified candidate variables to include, we conducted variable selection by random survival forest in the entire sample, while masked to treatment allocation. We used imputation for missing values in an algorithm to select covariates for inclusion in a TMLE model. Time-to-event analysis was computed from the time of transfusion until an endpoint. A one-sided test with type I error of 0.05 was used to determine statistical significance.

## RESULTS

### TRIAL POPULATION

From June 3, 2020 to October 1, 2021, a total of 1225 participants at 23 sites were randomized with 589 transfused with control plasma and 592 transfused with CCP for 1181 participants included in the mITT analysis (**Figure 1**). The trial was halted after enrolling over 90% of its initial target due to declining hospitalizations among enrolled participants (5-6 per hundred enrollees for the first 1000 enrollees, but less than 1 per hundred enrollees thereafter).

**Figure 1.**
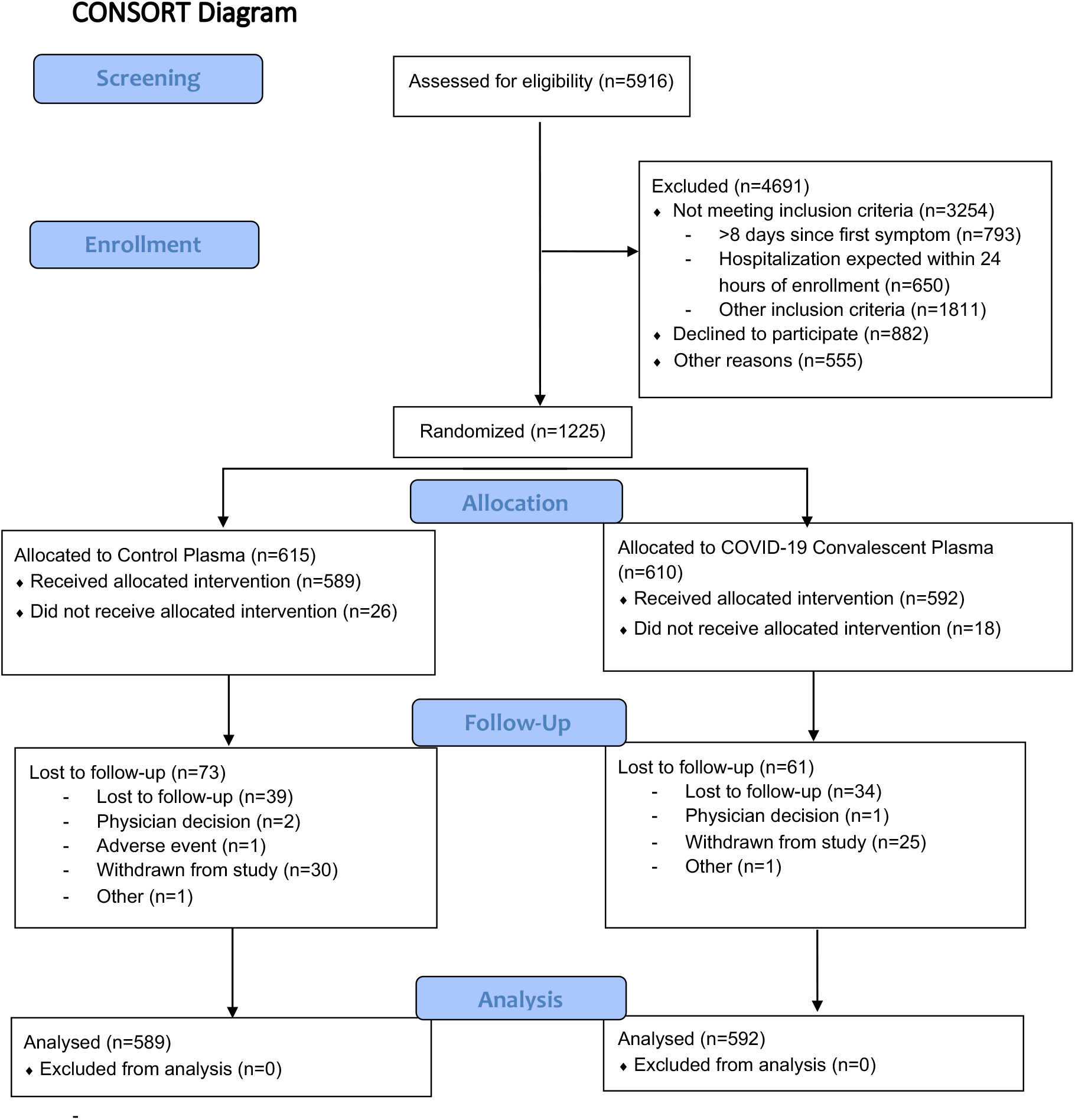
Enrollment, Randomization and Treatment Populations. Potential participants who a diagnostic test positive for SARS-CoV-2 and ≤ 8 days of COVID-19 were assessed both for eligibility by study personnel and by investigators to confirm that they were safe for outpatient management. Participants may have had ≥1 reason for exclusion from the trial. The intention-to-treat population included all randomized participants and the modified-ITT excluded randomized participants who did not receive assigned trial product.

There were no clinically significant imbalances in baseline characteristics, comorbidities, COVID-19 vaccines, vital signs or clinical laboratory results (**Table 1**). The median age was 44 years with 80 (7%) ≥65 years and 411 (35%) ≥ 50 years. Black/African American (n=163) or Hispanic (n=170) each comprised more than 10% of the participants. American Indian or Native Pacific Islander were 21 (2%) total. Women represented 57% of the participants, including three pregnant participants. The median time from symptom onset to transfusion was 6 days.

**Table 1:**
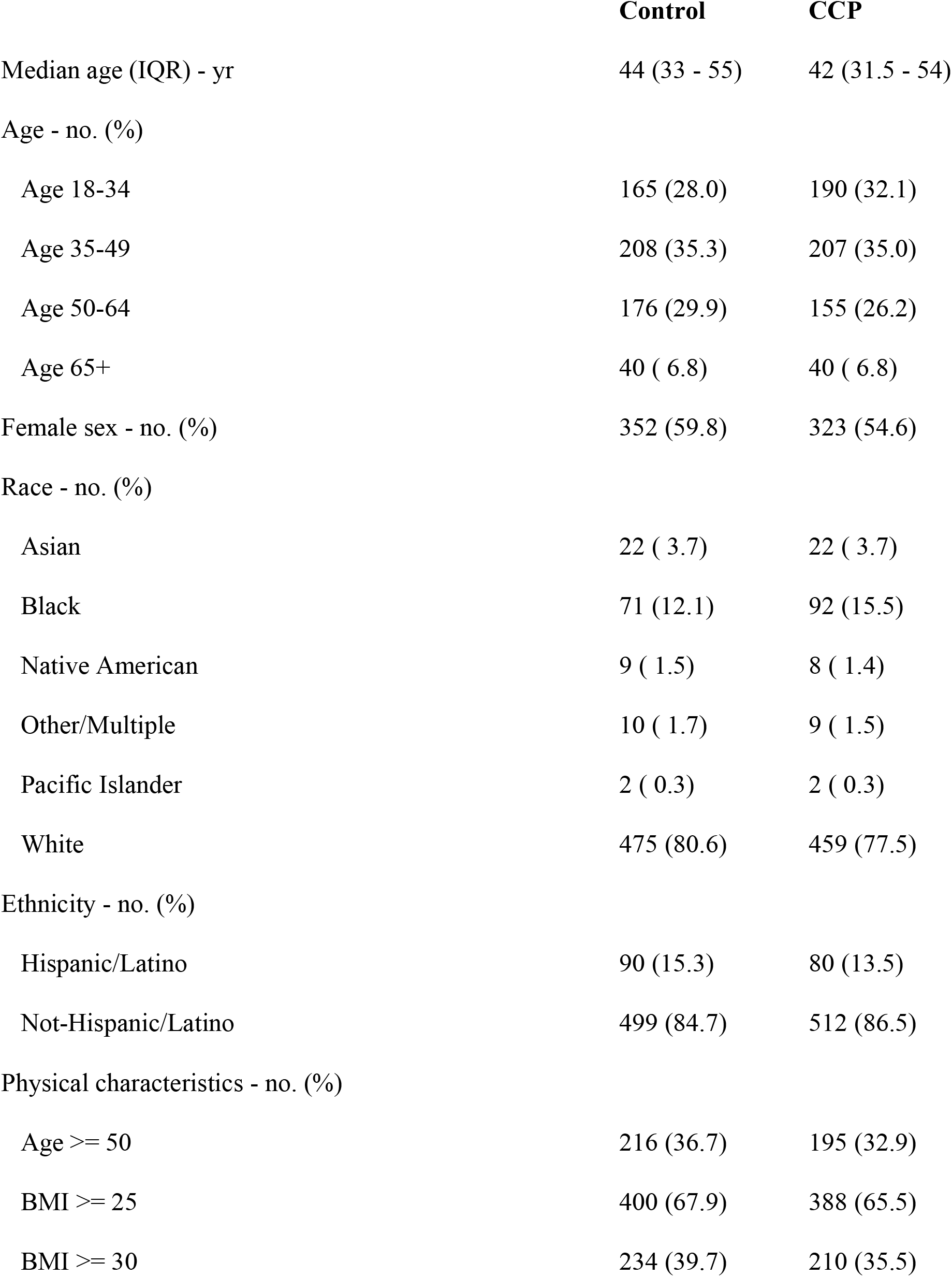
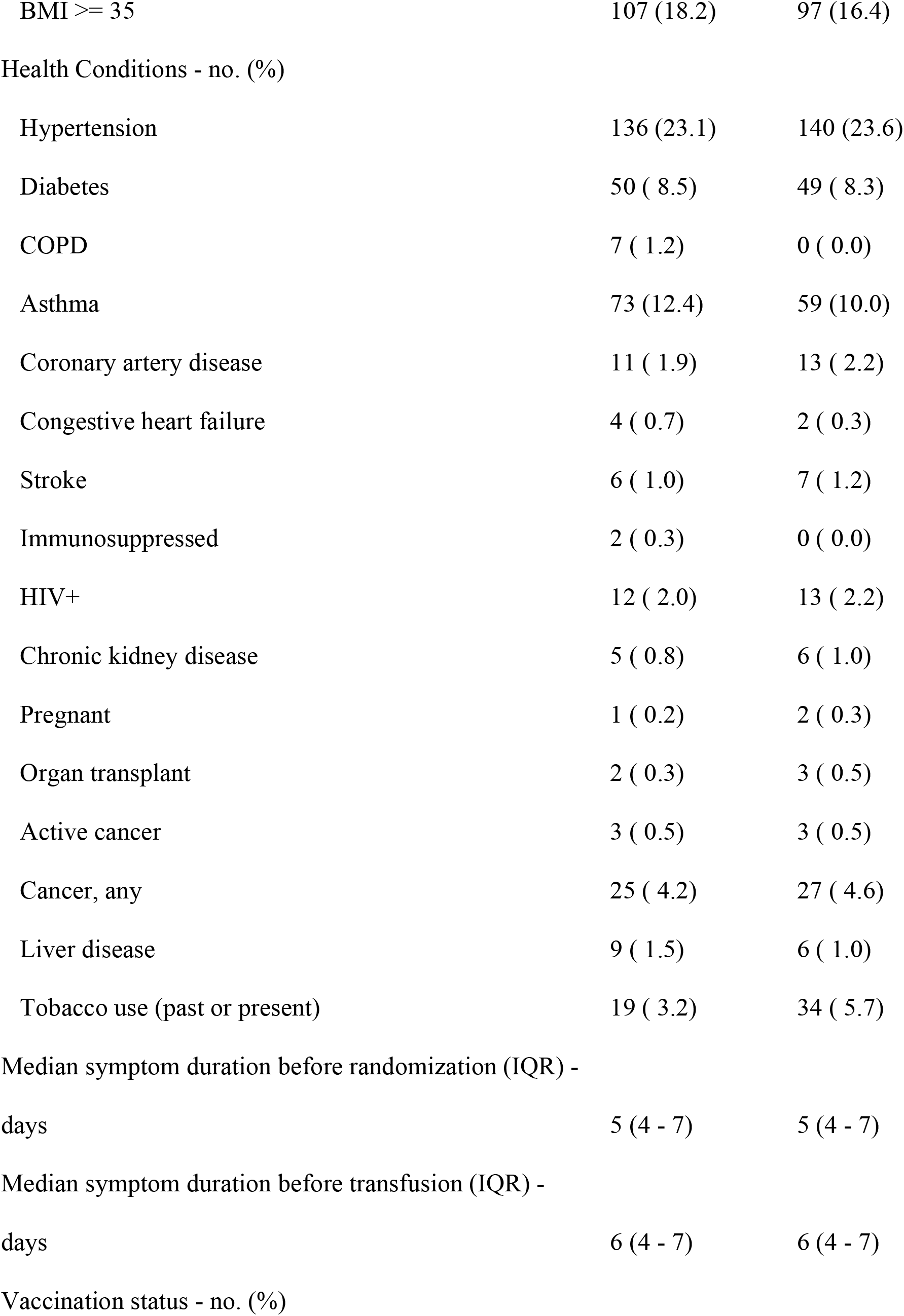
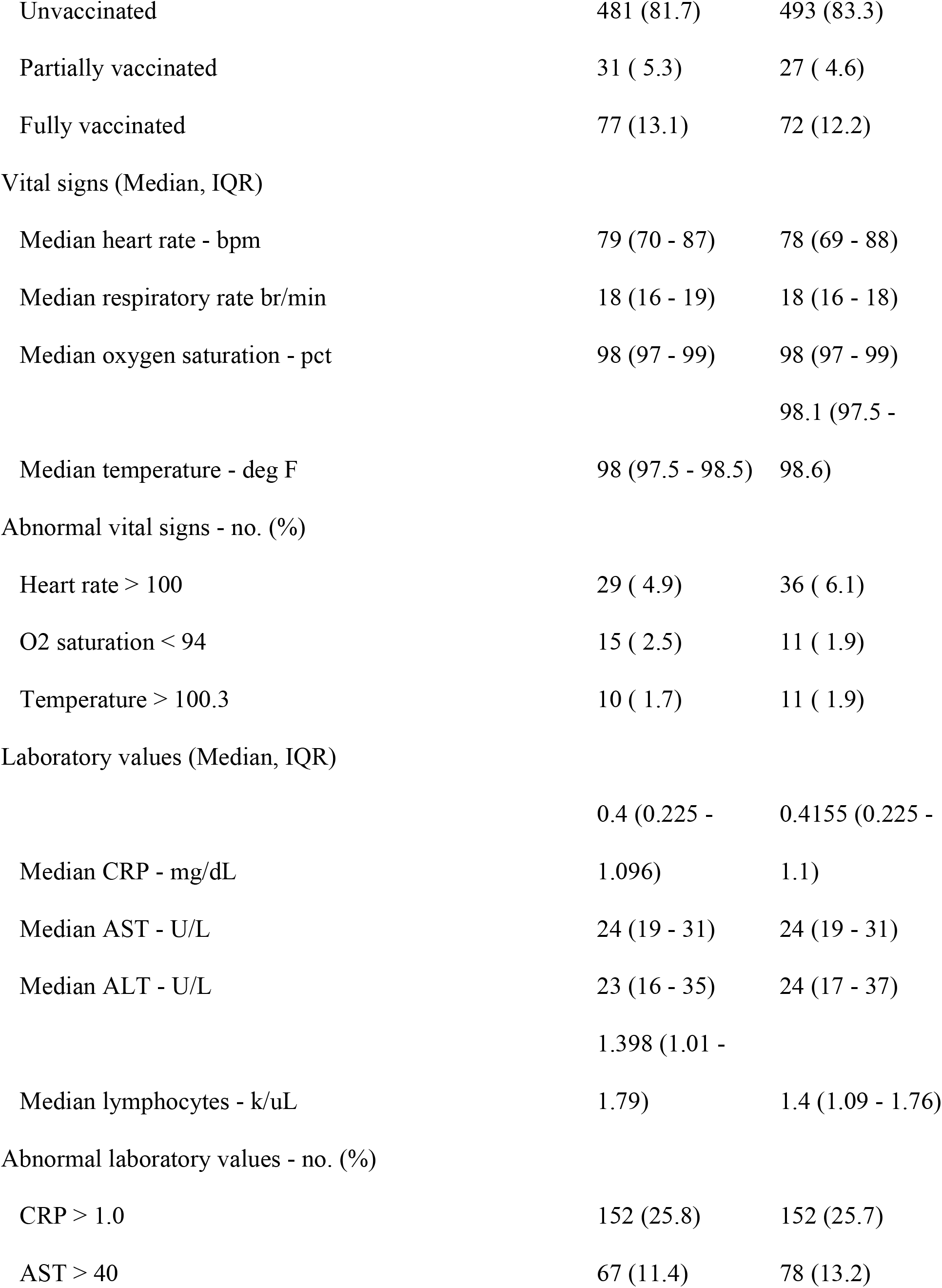
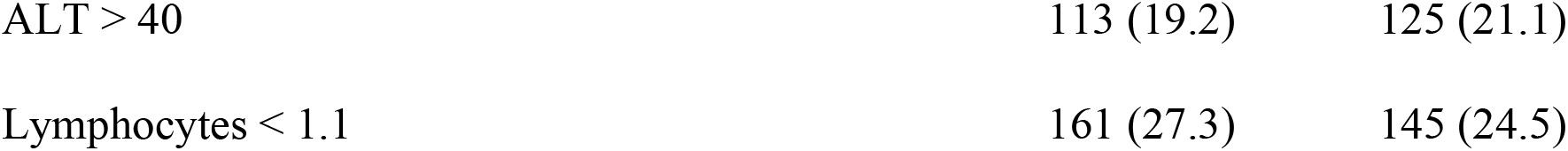
Baseline of mITT population characteristics by treatment arms.

|  | **Control** | **CCP** |
| --- | --- | --- |
| Median age (IQR) - yr | 44 (33 - 55) | 42 (31.5 - 54) |
| Age - no. (%) |  |  |
| Age 18-34 | 165 (28.0) | 190 (32.1) |
| Age 35-49 | 208 (35.3) | 207 (35.0) |
| Age 50-64 | 176 (29.9) | 155 (26.2) |
| Age 65+ | 40 ( 6.8) | 40 ( 6.8) |
| Female sex - no. (%) | 352 (59.8) | 323 (54.6) |
| Race - no. (%) |  |  |
| Asian | 22 ( 3.7) | 22 ( 3.7) |
| Black | 71 (12.1) | 92 (15.5) |
| Native American | 9 ( 1.5) | 8 ( 1.4) |
| Other/Multiple | 10 ( 1.7) | 9 ( 1.5) |
| Pacific Islander | 2 ( 0.3) | 2 ( 0.3) |
| White | 475 (80.6) | 459 (77.5) |
| Ethnicity - no. (%) |  |  |
| Hispanic/Latino | 90 (15.3) | 80 (13.5) |
| Not-Hispanic/Latino | 499 (84.7) | 512 (86.5) |
| Physical characteristics - no. (%) |  |  |
| Age >= 50 | 216 (36.7) | 195 (32.9) |
| BMI >= 25 | 400 (67.9) | 388 (65.5) |
| BMI >= 30 | 234 (39.7) | 210 (35.5) |
| BMI ≥ 35 | 107 (18.2) | 97 (16.4) |
| Health Conditions - no. (%) |  |  |
| Hypertension | 136 (23.1) | 140 (23.6) |
| Diabetes | 50 ( 8.5) | 49 ( 8.3) |
| COPD | 7 ( 1.2) | 0 ( 0.0) |
| Asthma | 73 (12.4) | 59 (10.0) |
| Coronary artery disease | 11 ( 1.9) | 13 ( 2.2) |
| Congestive heart failure | 4 ( 0.7) | 2 ( 0.3) |
| Stroke | 6 ( 1.0) | 7 ( 1.2) |
| Immunosuppressed | 2 ( 0.3) | 0 ( 0.0) |
| HIV+ | 12 ( 2.0) | 13 ( 2.2) |
| Chronic kidney disease | 5 ( 0.8) | 6 ( 1.0) |
| Pregnant | 1 ( 0.2) | 2 ( 0.3) |
| Organ transplant | 2 ( 0.3) | 3 ( 0.5) |
| Active cancer | 3 ( 0.5) | 3 ( 0.5) |
| Cancer, any | 25 ( 4.2) | 27 ( 4.6) |
| Liver disease | 9 ( 1.5) | 6 ( 1.0) |
| Tobacco use (past or present) | 19 ( 3.2) | 34 ( 5.7) |
| Median symptom duration before randomization (IQR) - |  |  |
| days | 5 (4 - 7) | 5 (4 - 7) |
| Median symptom duration before transfusion (IQR) - |  |  |
| days | 6 (4 - 7) | 6 (4 - 7) |
| Vaccination status - no. (%) |  |  |
| Unvaccinated | 481 (81.7) | 493 (83.3) |
| Partially vaccinated | 31 ( 5.3) | 27 ( 4.6) |
| Fully vaccinated | 77 (13.1) | 72 (12.2) |
| Vital signs (Median, IQR) |  |  |
| Median heart rate - bpm | 79 (70 - 87) | 78 (69 - 88) |
| Median respiratory rate br/min | 18 (16 - 19) | 18 (16 - 18) |
| Median oxygen saturation - pct | 98 (97 - 99) | 98 (97 - 99) |
|  |  | 98.1 (97.5 - |
| Median temperature - deg F | 98 (97.5 - 98.5) | 98.6) |
| Abnormal vital signs - no. (%) |  |  |
| Heart rate > 100 | 29 ( 4.9) | 36 ( 6.1) |
| O2 saturation < 94 | 15 ( 2.5) | 11 ( 1.9) |
| Temperature > 100.3 | 10 ( 1.7) | 11 ( 1.9) |
| Laboratory values (Median, IQR) |  |  |
|  | 0.4 (0.225 - | 0.4155 (0.225 - |
| Median CRP - mg/dL | 1.096) | 1.1) |
| Median AST - U/L | 24 (19 - 31) | 24 (19 - 31) |
| Median ALT - U/L | 23 (16 - 35) | 24 (17 - 37) |
|  | 1.398 (1.01 - |  |
| Median lymphocytes - k/uL | 1.79) | 1.4 (1.09 - 1.76) |
| Abnormal laboratory values - no. (%) |  |  |
| CRP > 1.0 | 152 (25.8) | 152 (25.7) |
| AST > 40 | 67 (11.4) | 78 (13.2) |
| ALT > 40 | 113 (19.2) | 125 (21.1) |
| Lymphocytes < 1.1 | 161 (27.3) | 145 (24.5) |

### STUDY CONVALESCENT PLASMA

There were 333 unique donor units transfused into 592 individuals with many identical plasma aliquots from large volume single donations going to 2-4 recipients. Of the CCP units 300 of 333 (90%) were donated from April to December 2020 with the remaining 33 in January to April of 2021. Plasma unit serological analysis revealed that 99% had SARS-CoV-2 spike protein antibody titers 1:1,620 or higher and 95% 1:4,820 or higher^20^. The unique plasma unit median titer was 1:14,580 with 81% meeting the current FDA high titer definition by Euroimmun testing^19^. Control plasma units were either donated in 2019 or tested seronegative for SARS-CoV-2.

### PRIMARY ENDPOINT-HOSPITALIZATIONS

In the prespecified mITT trial population excluding those not transfused, the endpoint COVID-19 related hospitalization within 28 days occurred in 37 of 589 participants who received placebo control plasma and in 17 of 592 participants who received CCP (relative risk [0.46] compared with controls; 95% upper limit confidence interval 0.733; P=0.004 (**Table 2**). The relative risk reduction was 54%. A pre-specified adjusted TMLE analysis yielded similar conclusions to the unadjusted analysis (**Table 2**) as the cumulative incidences were similar (**Figure 2a**), suggesting a longer time (one day) to (**Table 2, Figure 2b**) and a lower risk (4%) of (**Table 2, Figure 2c**) hospitalization among those randomized to CCP vs. control. Results were similar in subgroups defined by sex and vaccination status (**Table 2**). The hospitalizations occurred predominately in unvaccinated individuals (53/54). The antibody levels in the CCP units transfused into the 17 subsequent hospitalized participants were similar to antibody levels in the CCP units transfused into those not hospitalized. The duration of hospitalization was similar at 6 days in both groups after excluding the three deaths in the control group.

**Table 2.** COVID-19 hospitalization or death prior to day 28 by treatment group.

|  | **Control** | **CCP** |  |
| --- | --- | --- | --- |
|  | **(N=589)** | **(N=592)** | **P-value<sup>1</sup>** |
| **Number randomized, n** | **615** | **610** |  |
| **Number transfused, n** | **589** | **592** |  |
| **Participants with primary outcome** | **37** | **17** | **0.004** |
| **(hospitalization), n** |  |  |  |
| Unadjusted Relative Risk [95% one-sided CI upper bound] |  | 0.46 [0.73] | 0.004 |
| **Hospitalization not due to COVID-19<sup>2</sup>, n** | **3** | **4** | **0.77** |
| **Details of primary outcome severity, n** |  |  |  |
| Death after hospitalization | 3 | 0 |  |
| ICU Hospitalization (no mechanical ventilation) | 4 | 3 |  |
| Non-ICU hospitalization due to COVID-19, requiring supplemental oxygen | 26 | 12 |  |
| Non-ICU hospitalization due to COVID-19, not requiring supplemental oxygen | 4 | 2 |  |
| A stay of >24 hours for observation in an ED, field hospital or other healthcare unit or receipt of oxygen for >24 hours, outside of hospital | 0 | 0 |  |

<sup>1</sup> P-values calculated using one-sided Fisher's exact test for count data and Cochran-Mantel-Haenszel test.
<sup>2</sup> Number of first hospitalizations that were not due to COVID-19 (i.e.- hallucinations due to pre-existing mental illness, renal colic, constipation or pancreatitis)

|  |  |  |  |
| --- | --- | --- | --- |
| **Expected time free of primary outcome<sup>3</sup> (days)** | **26.27** | **27.26** |  |
|  |  | 0.99 (0.28) |  |
| Difference (SE) [95% one-sided CI lower bound] |  | [0.54] | 0.0002 |
| **Probability of remaining free of hospitalization<sup>3</sup>** | **0.93** | **0.97** |  |
|  |  | -0.04 |  |
| Difference (SE) [95% one-sided CI lower bound] |  | (0.01)[-0.02] | 0.0003 |
| **Primary outcome events by sex<sup>4</sup>** |  |  |  |
| Female | 21 | 9 |  |
| Male | 16 | 8 |  |
| **Primary outcome events by vaccination status<sup>4,5</sup>** |  |  |  |
| Unvaccinated | 36 | 17 |  |
| Partially vaccinated | 1 | 0 |  |
| Fully vaccinated | 0 | 0 |  |

<sup>3</sup> Adjusted for age, trial site, BMI, baseline albumin, bicarbonate, c-reactive protein, glucose, potassium, and baseline abnormal head, eyes, ears, nose, and throat physical examination, as specified from a random survival forest analysis of baseline characteristics. Primary analysis data set restricted via principal component analysis, reducing data set to 990 complete case participants with full data (494 transfused with control plasma, 496 transfused with convalescent plasma). Changing the model by decreasing covariates did not change inferences.
<sup>4</sup> Numbers of participants in each category found on Table 1
<sup>5</sup> Fully vaccinated 14 days following final administration
<sup>6</sup> Cox proportional hazards models with only treatment assignment and then adjusted for age; age and site; age site, and BMI; and finally same covariates as TMLE model, showed congruence with TMLE results as hazard ratios were approximately 0.45 across proportional hazards models.

**Figure 2.**
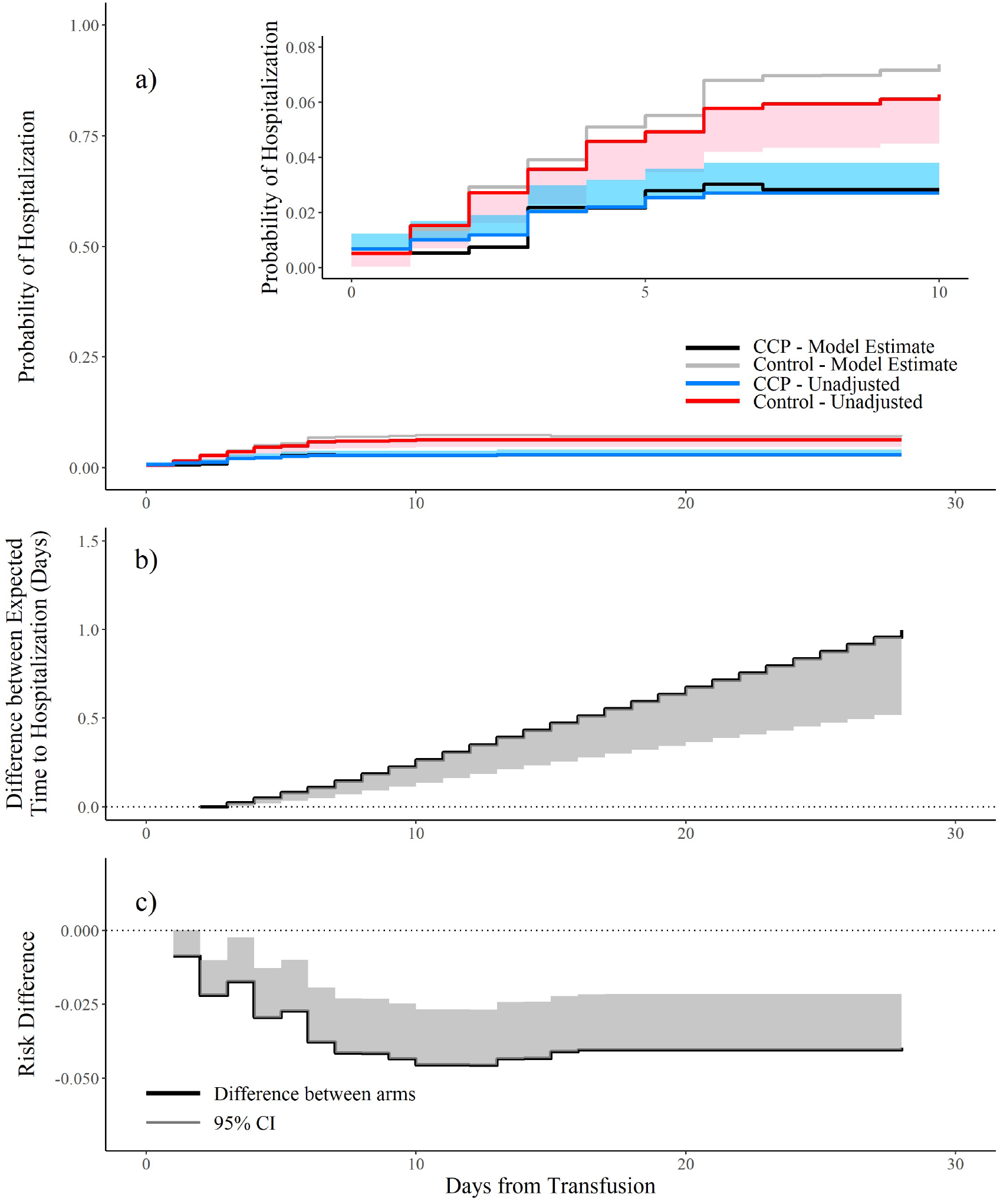
Probability of hospitalization. a). Cumulative incidence of COVID-19 related hospitalization, unadjusted with confidence interval presented along with TMLE model estimates; b). adjusted estimated difference in the expected time to hospitalization (≥ 0: increased expected days to hospitalization for CCP); c). adjusted estimate of the risk difference between treatment arms (<0: lower risk of hospitalization for CCP). 95% CI=One-sided 95% confidence interval.

### HOSPITAL SEVERITY

Among the mITT population 26 in the control group and 12 participants in the CCP group progressed to a hospital oxygen requirement (**Table 2**). All 3 post-hospitalization deaths were in the control plasma group.

### OTHER TRIAL ANALYSES

In an intention-to treat analysis including all randomizations (1225) and all participants with hospitalizations (64) within 28 days of transfusion, hospitalizations occurred in 42 of 615 (6.8%) randomized control participants and in 22 of 610 (3.8%) CCP participants (relative risk 0.52; 95% upper bound confidence interval 0.806; P=0.008) (**Table 2**). Three hospitalized individuals were consented, randomized but not transfused, and 7 hospitalizations were adjudicated as non-COVID-19 related hospitalizations prior to unblinding. All 1181 transfused participants had confirmed hospital status by day 28.

### SAFETY

There were 87 total reported AEs, 53 in the control group and 34 in the CCP group (**Table S1**). All 44 of the listed AE pneumonias were also included in the Table 2 hospitalizations. One transfusion was stopped after 2-3 mL were administered, secondary to development of diffuse erythema and nausea; the participant was evaluated in the ED and discharged (Table S2). One control group individual progressed to acute respiratory distress syndrome (ARDS), mechanical ventilation and death adjudicated secondary to COVID-19 (**Table S3**).

## DISCUSSION

In this randomized trial, administration of high titer SARS-CoV-2 convalescent plasma reduced outpatient hospitalizations by more than 50% among participants with recent infection. Immune sera or plasma has been used safely for infectious diseases treatment for over 100 years^16^. Mixed results in previous infectious diseases outbreaks may be due to lack of modern study designs, small sample sizes, differential viral response to passive antibodies, inclusion of low titer antibody units, or late administration in relation to disease onset^22^. The results of our multi-site study strengthen prior experience with antibody-based therapies, which show effectiveness requires early administration with sufficient quantity to mediate an antiviral effect^22^.

Our study builds upon and enlarges the impact of the Argentinian study in 160 older COVID-19 outpatients, randomized to received CCP or placebo within three days of symptom onset, which demonstrated a 48% reduced hypoxia or tachypnea risk ^17^. Our findings in all age groups with 75% transfused beyond 3 days may be more practical due to implementation delays in diagnostic testing. Our study results stand in contrast to the CCP COVID-19 treatment study, conducted at 48 EDs^18^. Patient enrollment during ED presentation, possibly represents a population with more advanced disease. Indeed, a quarter of the hospitalized patients met the endpoint during the ED randomization/transfusion visit, limiting the time for CCP to exert an effect. Additionally, the trial outcome included an equal number of return visits to the ED/urgent care in the treatment arms.

Likewise, our findings are similar to those of trials that evaluated mAbs to SARS-CoV-2 with regards to efficacy including the magnitude of effect (50 to 70% reduction in medical visits)^1^. Our study included a participant population with symptoms up to 8 days, whereas the sotrovimab trials included symptoms less than 5 days^2^ and the bamlanivumab and etesevimab study were limited to 3 days^3^. In both interventions, the active agent is viral specific antibody. Whereas mAbs are available in high-income countries, they are expensive to produce, require time for new drug approval and may not be widely available during surge conditions. In contrast, CCP is available in low- and middle-income countries, has no patent limitations, and is relatively inexpensive to produce, with many single donors being able to provide multiple high titer units, as evident from this trial. Because it provides a diverse mix of antibodies with different specificities and functions, CCP is much less vulnerable to the emergence of antibody resistance In fact, CCP has been used for rescue therapy in immunocompromised patients who developed mAb-resistant SARS-CoV-2 variants^5^. Since any individual who recovers from variant SARS-CoV-2 mounts antibodies against that variant, CCP is an antibody-based therapy that locally keeps up with variants^23^. Hence, CCP is likely to remain an important therapeutic option for COVID-19.

In our study, the most common reason for hospitalization was symptomatic hypoxia, resulting from pulmonary inflammation in response to SARS-CoV-2 infection. Plasma antibodies mediate several antiviral activities including direct virus neutralization, complement activation, viral particle phagocytosis and antibody-dependent cellular cytotoxicity^24^. The accumulated COVID-19 vaccine data point to lower antibody levels to prevent severe disease than infection^25^.

Our study faced important challenges. First, standards of care and available therapies changed throughout the study period. Anti-SARS-CoV-2 mAbs became available in late November 2020, which steadily decreased the individuals eligible for CCP. Also, as vaccine utilization increased, the frequency of study hospitalizations decreased. The variants of concern became more prevalent during the study period, first with alpha and then delta in summer of 2021. The study plasma was largely obtained in 2020 from donors who recovered from COVID-19 with ancestral forms of SARS-CoV-2. The trial logistics required multiple blood banks with the ability to provide plasma for all blood types at 23 sites during a pandemic when many health care systems were working on limited, fluctuating capacity. However, routine blood banking standards were able to support proper supply logistics with remote coordination. The risk of SARS-CoV-2 infection required appropriate infection prevention measures in outpatient sites, often specially constructed and separated from hospital populations. The safety of CCP allowed for pregnant women to enroll, a population at high risk for COVID-19 progression who have been excluded from previous COVID-19 treatment trials.

In addition to challenges, our trial had limitations. First, for practical purposes, the trial outcome was hospitalization for COVID-19, not deaths. Noteworthy, the three participant deaths occurred in the control plasma group. Second, the hospitalized rate in the control group was 6%, less than the 8% average for the United States. Third, only 35% of participants transfused were over age 50 and the trial was not large enough for definitive subgroup analyses on medical comorbidities or pregnancy. Strengths of this randomized controlled trial include a large, diverse study population enrolled at over 20 sites throughout the United States, inclusion of all ages over 18 years, double blind intervention with control plasma and high rates of both transfusion and follow-up.

Our trial has important public health implications, especially in resource-constrained settings. High titer CCP should be considered for initial deployment in COVID-19 and future pandemics while monoclonal therapies and vaccines are being developed. This simple and potentially inexpensive intervention may reduce early pandemic morbidity and mortality. Development of infusion centers that can rapidly deploy CCP for outpatient pandemic use will be an important consideration for future health care systems. Even in the current pandemic, continued propagation of SARS-CoV-2 variants with evolving resistance to currently available antivirals and even mAbs, should raise the importance of maintaining capacity to rapidly escalate CCP availability and distribution especially as locally sourced recent plasma should include antibodies to circulating strains^26^. Antibody concentration is heterogeneous among donors^20^ and future pandemics should consider restricting therapeutic plasma to the upper 30 to 40 percent of plasma units with high titer over 1:1000.

In conclusion, early high titer SARS-CoV-2 convalescent plasma outpatient administration reduced hospitalizations by more than 50%. Given our robust findings and concordance with the small Argentinian ambulatory trial^17^, high titer CCP should be considered for outpatient COVID-19 care as an extension of current hospital based CCP EUA.

## Data Availability

All data produced in the present study are available upon reasonable request to the authors

## Acknowledgments

The authors gratefully acknowledge the study participants who generously gave of their time and biological specimens. Initial work was catalyzed by grants from Bloomberg Philanthropies and the State of Maryland

