## Appendix for "Randomized Controlled Trial of Early Outpatient COVID-19 Treatment with High-Titer Convalescent Plasma"

Supplementary Appendix

This appendix has been provided by the authors to give readers additional information about their work.

Supplement to: Sullivan, Gebo Shoham et al

Table of Contents


| | |
|---|---|
| Enrolling Centers, Principal Investigators | 2 |
| Blood Bank Staff | 7 |
| CSSC Leadership Staff | 10 |
| Summary of protocol changes | 12 |
| Supplement Table 1 | 14 |
| Supplement Table 2 | 16 |
| Supplement Table 3 | 17 |

**Enrolling Centers Principal Investigators (in order of enrollment contributions)**

Johns Hopkins University (437) CoPIs- David Sullivan, Kelly Gebo, Shmuel Shoham

Jeanne Keruly, Diane Lanham, Sonali Thapa, Smita Khati, Atika Singh, Bukola Adeosun, Amrita Raj, Eugene Bosworth, Jessica Coleman, Stephanie Katz, Kim Kafka, MiKaela Olsen, Cynthia Sparks, Ann Folmer, Belinda Overstreet, Vicky Virina, Bilenda Vazaian, Angie VanTassel, Joanne Harkness, Jennifer Clinnin, Shelby Brumback, Amanda Pennes, Sherilyn Brinkley

MedStar Washington (110) - PI-Aarthi Shenoy

Patricia Tanjutco, Theresa M. Moriarty, Kristin Garman, Sandra Griffin, Maureen McNulty, Laira Baez-Quintana

NorthShore University (88)- PI-Giselle Mosnaim

Amanda Caplan, Alvin Thomas, Amanpreet Kalkat, Thomas Gniadek, Ewa Schafer, Leonard Kaplan, Rachel Story, Shashi Bellam, Thomas Hensing, Tracy Spizzirri, Giselle Martinez-O'Connor, Ruvini De Vas Gunawardhane, Marycatherine Devitt, Justin Brueck, Robert Stanton, Margo Quinn, Rachel Pulido, Darryck Mauer, Brittney Adams-Crutcher, Candy Gonzalez, Patricia Debosz, Julie Anderson, Milena Draganov, Lisa Akelian

Baylor College of Medicine (81) - PI-Yuriko Fukuta

Ahmed Hamdi, Christopher Amos, Galant Chan, Tyler Lambing, Zohra Huda, Eva Zimmerman, Crystal Diaz, Ryan Mathew, Anne McCombs, Carrie Kibler, Leah Doyle, Samantha Macias, Abdullah Esmail, Rene Plascencia, Alicia Lawson, Lourdes Garcia Pelaez

Univ Texas, Houston (75) - PI-Bela Patel

Reeba Mathew, Lilit Sargsyan, Ruckshanda Majid, Isabel Mira-Avendano, Mary Ayad, Luis Ostrosky, Henry Wang, Farah Akhtar, Daniel Hwang, Tanya Khan, Pamela Nichols, Jaicey

Johnson, Mary Rangel, Iram Abbassi, Trini Tran, Marie Cruz, Lopez Fallon, Greg Killer, Sandra Garcai, Cursheena Stanciel, Rhonda Hobbs, Maria Hernandez, Carolyn Grimes,

University of Alabama at Birmingham (71) - PI-Sonya Heath

Andrew Yousef, Anthwen Washington, Michael Fordham, Tommy Williams, Michelle Chambers, Faye Heard, Jasmine McQueen, Charlotte Olivia Hogue, Dorothy Nieters, Susan Ellen Binkley, Thomas Patrick Frazier, Deanna Chandler, Elizabeth Valentine, Bianca Gooden, Alisha Hitt, April Mack, Edgar Overton, Todd McCarty, Randall Davis

Rhode Island Hospital/Brown University (67) - PI-Adam C. Levine

Adam Aluisio, Alexis Lawrence, Alison Kelley, Alyssa Peachey, Amanda Block, Ahsley Gaipo, Austin Lee, Cheri Antunes, Christine Penfield, Christopher Shakespeare, Esabella Cesarini, Evaniz Suarez, Francesca Beaudoin, Frank Overly, Gregory Jay, Huy Nguyen, Jamie Chapman, Jeffrey Bailey, Jhanavi Kapadia, Jheraldines Paulino, Katelyn Moretti, Kristin Basso, Margaret Geoghegan, Lindsay McKeever, Lisa Tingley, Matthew Shiavo, Naz Karim, Ramu Kharel, Samuel Kaplan, Sarah Ducharme, Sarah Tokarz, Selim Suner, Sonya Naganathan, Taneisha Wilson, Tarek Nizami, Weston DeLomba, William Binder, Yanira DePina

Luminis Health-Anne Arundel Med Center (37)- PI-Barry Meisenberg

Venita Alston Crawford, Erika Siegrist, Emily Ross, Jaci Miller, Lun Chen, Dmitriy Pinelis, Clark Johnson, Susan Crawford, Michael Thompson, Olumide Ayeni, Sarah Licari, Sally Keckeisen, Marie Tangu

University of Utah (36) - PI-Emily Spivak

J. Robert Singleton, Evan Heller, Kasra Rahmati, Carson Espinoza, Lloyd Stanley Pies, Josh Fife, Michelle Adams, Dee Ann Ashby, Shawna Baker, Pam Dansie, Jesika England, Samantha

Hess, Jenna Lumpkin, Kathy Lusted, Virginia Mast, Jonah Simmons, Kelsey Webb, Ryane Sorensen, Kimberly Rhodes, Dixie Thompson, Lucas Cavazotti

University of Miami (35) - PI-Shweta Anjan

Yanyun Wu, Lilian Abbo, Jose Gonzales Zamora, Laura Beauchamps, Jovanna Betran-Lopez, Emilia Faraj, Maria Almanzar, Claudia Bornia, Veronica Del Prete, Diana Saez, Benita Abatayo,

University of Cincinnati (27) - PI-Moises Huaman

Carl Fichtenbaum, David Oh, Linda Hinds, Eva Whitehead, Jennifer Baer, Sarah Trentman, Sharon Kohrs, Michele O'Neill, Lyndsey Armor, Brenda Miller, Helen Shelton, Anissa Moussa, Marlena Petrie, D'Vaughn House

Mayo Clinic Arizona (24)-PI-Janis Blair

Jill Adamski, Emily Frank, Tahnie Taylor, Amy Kole, Mark Burns

UCLA (23)- PI-Judith Currier

Paul Allyn, Omer Beaird, Marianna Bernando, Claire Brown, Meilani Cayabyab, Kara Chew, Jesse Clark, Rafael Corona, Madeline Cross, Jaime Deville, Anaar Eastoak-Siletz, Noah Federman, Jennifer Fulcher, Risa Hoffman, Nicholas Hornstein, Paul Krogstad, Raphael Landovitz, Nancy Lopez, Lisa Mark, Andrea McGonigle, Zhen Mei, Arash Naeim, Kyron Pierce, Kavitha Prabaker, Adreanne Rivera, Sarahmay Sanchez, Melanie Shmitz, Max Schumm, Areti Tillou, John Timmerman, Tran Tran, Stefani Uechi, Tara Vijayan

Wayne State University (22) - PI-James Paxton

Brian O'Neil, Robert Welch, Samuel Ceckowski, Theodore Falcon, Sondall Hulin, Thomas Mazzocco, Justin Sabol

University of Mass, Worcester (21) - PI-Jonathan Gerber

Caitlin Pasquale, Tzafra Tessier, Lisa Ford, Ryan Horrgan, Lindsey Shanahan, Tara Roy

Nuvance Health- Joanne Petrini, William Rausch, Marie Elena Cordisco. Jon Serino, Bryan Lewis, Daniel Cruser

Nuvance Health Danbury Hospital (19) - PI-Patrick Broderick

Reed Idriss, Brian McCambley, Robert Bazuro, Douglas Smith, Tammy Weiner, Michael Emond, Melissa Saad, Teresa Pearson, Bhumi Shah, Nusrat Pathan, Svetlana O'Connor, Christine Oliveira, Dawn Marie Morsey

Nuvance Health Vassar Brothers Medical Center (9)- PI-Valerie Cluzet

BethAnne Altier, Catherine Attanasio, Barakat Onabanjo, Tricia Landi, Carissa A Sharp, Camille Finkle, John Haumaier, Susan Chmura MT(ASCP), Nicole Atkins, Charlotte Murray, Marie Bien-Aime, Tuyen Nguyen

Nuvance Health Norwalk Hospital (1) - PI-Jean Hammel, Benjamin Greenblatt

Elizabeth Luttner,Rebecca Martinez, Jose Leonel Rodriquez, Jose Leonel Rodriquez Milisha Patel, Norman Jones MT (ASCP), Michael Camussi, Mary Pattison,

Ascada Research (19) PI-Kevin Oei, Matthew Abinante

Gaurav Thakral, Nawar Hussin, Kevin Hoang, Pearl Chen, Alyssa Holt, Nathan Tauta, Marvy Abraham, Shawna Galbreath, Elizabeth Magana, Ghada Saleh, Rosalina Hernandez, Yasmine Retana, Desiree Fimbres, Denise Solia

JHSPH Center for American Indian Health (12)- PI-Laura Hammitt

Laura Brown, Hanna Foster, Kristen Roessler, Catherine Sutcliffe, Robert Weatherholtz, Natalie Jones, Angelina Reid, Diana Quay, Janene Colelay, Felicia Riley, Courtney Parkinson, Rebecca Larsen, Dennie Parker, Megan Gardner, Melinda Charley, Monique Harvey, Ella Paul, Melissa Christianson, Susie Dixon

UC Irvine (7)- PI-Donald Forthal

Minh-Ha Tran, Anthony Warner, Rosie Magallon, Nancy Hernandez, Jon Fox

University of Rochester (3) - PI-Martin Zand

Neal Blumberg, Omar Ajitali, Omar Aljitawi, David DeWolfe, Ann Miller, Susanne Heininger, Debbie Mase, Aimee, Diane Bullock, Sarah Magri, Christopher Lane

University of New Mexico (1 transfer)-PI-Jay Raval

Justin Baca, Marija Zimkute, Rebecca Brito, Christine Ferguson, Dominique Spence, Sarah Lavelle, Sandra Cano, Silas Bussmann, M. Nichole Trujillo

**Blood Banking staff**

Johns Hopkins Hospital

Christi Marshall MT(ASCP)BB, CQA(ASQ), Rivcah Davis MT, Melissa Neally MSHM, MLS(ASCP)CM SBB(ASCP), Kristen Buban MLS(ASCP)CM SBBCM

Luminis Health Anne Arundel Medical Center

Sanford Robbins M.D., Megan Frisk M.S., MLS (ASCP) CM SBBCM

Ascada Research Hoag Blood Bank

Arell Shapiro M.D. and Daniel Pach MT(ASCP), SBB, MBA

Baylor College of Medicine

Meredith Reyes, MD, Laura Korte, MBA,MT(ASCP)SBB, CCRP, Patricia Guerrettaz and Mei Pang Nguyen

JHSPH Center for American Indian Health

Thomas Stephens, CRN MPH, Joseph Fowler, CRN, Kim Taylor, CRN

Lifespan/Brown University: Rhode Island Hospital

Lisa Tingley, Tracey Cheves and Karen King

MedStar Washington Hospital Center

Kathleen (Cathy) Cantilena M.D, Lorraine Wyne MT

NorthShore University Health System

Thomas J. Gniadek M.D. Ph.D., Jessica Mallek, Susan Alexander, Gregory Wright MT(ASCP) SBB, CQA(ASQ)

Mayo Clinic

Jill Adamski, M.D., Ph.D.

UCLA

Alyssa Ziman M.D, Bridget Wallace MT(ASCP), SBB, Andrea McGonigle, Zhen Mei, Dawn Ward

University of Alabama at Birmingham

Marisa Marques M.D, Tammy Gray CLS, Ashton Kornbrust, Sarah Herring, Benjamin Lenard

University of California, Irvine

Minh-Ha Tran M.D, Belinda Kluchnikov MT, Marcela Feria II, Faten Mahmoud, Melissa Morris

University of California, San Diego

Angela Varela MT (ASCP), CLS, Maria (Pia) Barron, Dominga Espinoza

University of Cincinnati Medical Center

David OH, M.D. and Laura Demartino MT(ASCP) SBBcm

University of Massachusetts Worcester

Yong (David) Zhao M.D. and Natalie Malvick

University of Miami-Jackson Memorial Hospital

Yanyun Wu M.D., Mario Charles MT

University of New Mexico

Eileen Sierra, BSMT (ASCP) and Cindy Jones, BSMT (ASCP)

University of Rochester

Neil Blumberg M.D, Debra Masel MT, Diane Bullock, Sarah Magri, Aimee Kievitt

University of Texas Health Science Center at Houston

Rhonda Hobbs BS MT(ASCP)SBB and Veronica Alvarado

University of Utah Health

Kelly Cali MT CLS, Ryan Metcalf, Yazmin Barrera, Candace Thomas, Audrey Wise, Cassidy Rapier, Cody Sarle

Vassar Brothers Medical Center

Daniel Cruser M.D, Susan Chumura MT(ASCP), Nicole Atkins, Charlotte Murray, Marie Bien-Aime, Tuyen Nguyen

Wayne State University

Tammon Nash, MD, Sondall Hulin MT, Jim Fiedor, Sophia Hadjiev,

Nuvance Health Vassar Brothers Medical Center

Daniel Cruser M.D, Susan Chmura MT(ASCP), Nicole Atkins, Charlotte Murray, Marie Bien-Aime, Tuyen Nguyen

Nuvance Health Danbury Hospital

Nusrat Pathan M.D, Theresa Pearson MT , Bhumi Shah , Svetlana O'Connor

Nuvance Health Norwalk Hospital

Bhavna Khandpur M.D, Rebecca Martinez, Jose Leonel Rodriquez, Jose Leonel Rodriquez

Milisha Patel, Norman Jones MT (ASCP)

The CSSC leadership

David Sullivan MD, Kelly Gebo MD, Shmuel Shoham MD, Evan Bloch MD, PhD, Arturo Casadevall MD, PhD, Aaron Tobian MD and Daniel Hanley MD

Clinical Coordinating Center Leadership

Daniel Hanley, Daniel Ford, Karen Lane, Nichol McBee, Amy Gawad, Nicky Karlen, Erica Hopkins, Anusha Yarava, Lisa Walborn (MMI BSPH)

- Regulatory Core

  Nichol McBee, Daniel Amirault, Jordyn Carll

- Data Core

  Ying Wang, Piyali Das, Malathi Ram, Craig Ou

- QA Monitoring Core

  ACTALENT: Erica Hopkins, Kathy Bauer, Jessica Becker, Marc Buffaloe, Cheryl Carmody, Savario Cimini, Sonja Cooper, Omar Cummings, Zhenya Dozier, Eric Easter, Ann Glasse, Sonya Griffin, Erica Hopkins, Marjorie Hyderkhan, Wendy Jade, Suzanne Kensington, Aaron Klaevemann, Teresa Kvachuk, Julie Lacy, Lea Ma, Steven Malahias, Ann Marie Polito, Melanie Morabito, Steven Pittman, Christine Sears, Michelle Shrivers, Robyn Stuckey, Jennifer Young

  Emissary International, LLC: Steve Mayo, David Reichert, Nicky Karlen, Emily Blair, Sarah Lennington, Carolyn Koenig

- Investigational Plasma Core

  Karen Lane, Tom Gniadek, Christi Marshall, Anusha Yarava, Aaron Ye, Preeti Khanal, David Reichert

- Safety Core

Tracey Economas, Jordyn Carll

- Medical Monitor Core

  Ronald Rodriguez, MD., PhD, University of Texas Health Science Center at San Antonio

- Clinical Endpoint Committee

  Ronald Rodriguez, MD., PhD (Chair), University of Texas Health Science Center at San Antonio, Johanna Daily, MD, MS, Albert Einstein College of Medicine,

  Panagis Galiatsatos, MD, MHS, Johns Hopkins University

Data Coordinating Center

Bryan Lau, Stephan Ehrhardt, Dave Shade, Molly Duan and Sheriza Baksh

Data Safety Monitoring Board

Pablo Tebas University of Pennsylvania (as DSMB Chair),

Roy F. Chemaly, MD Anderson Cancer Center,

Joe Massaro, Boston University School of Public Health,

Keith Kaye, University of Michigan, Maya McKean-Peraza, Duke Clinical Research Institute (Administrator)

JHU Research Laboratory

Evan Bloch, Aaron Tobian, Andy Pekosz, Sabra Klein, Oliver Laeyendecker, Mario Caturegli, David Sullivan, Reinaldo Fernandez, Evan Beck, Janna Shapiro, Han-Sol Park, Christopher Caputo, Maggie Li, Ioannis Sitaris, Anne Jedlicka, Amanda Dziedzic Manju Thakar, Brett Dietz, Isaiah Hoffland, Yolanda Eby, Xianming Zhu, Olivia Akinde, Ruchee Shrestha, Annie Wu

The Bliss Group and The Next Practice - Clinical Trial Recruitment Acceleration and Diversity Specialists The Bliss Group - Michael Roth, Quintin Maidment, Meg Wildrick

The Next Practice - Colin Foster, Desiree Huitt, David Pierpont

**Summary of protocol changes**

**Inclusion/Exclusion**

1. Removed receipt of previous blood products as holdover from merged donor and recipient protocols

2. Any positive molecular test including saliva will qualify for study. PCR test is no longer sole means of qualified test.

3. Excluded participants with prior to enrollment drug receipt with established viral activity against virus.

4. Steroid treatment does not influence eligibility.

5. Receipt of vaccine prior to enrollment also does not exclude participation

6. Prior monoclonal receipt before study plasma excludes participation. However, study participants who choose may receive monoclonals after study plasma based upon availability.

**Study plasma product**

1. Qualified plasma at greater than 1:320 titer only without FDA current standard which did not exist in 2020.

2. Control plasma collected after Dec 31, 2019 will be tested seronegative for SARS-CoV-2 antibody.

3. Added minimum infusion volume of 175 mL with no maximum for both convalescent and control plasma.

4. After July 2021, convalescent plasma used in trial must also meet FDA criteria for high titer plasma in addition to greater than 1:320 titer.

**Statistical analytic plan**

1. Modified to one sided Type 1 error for superiority

2. Added futility evaluation by DSMB at 40% enrollment after 28 day visit.

3. Added TMLE to statistical analysis plan

4. Target of older age group recruitment was made “not binding” because of vaccines and monoclonals restricting older ages in study.

5. Detailed severity score of COVID-19

**Protocol implementation**

1. Added visit study windows to define time frame for assessment.

2. Clarified that full range of medical rescue therapy after reaching hospital endpoint with full participation in rest of protocol. Participants remain blinded during hospitalization and for remaining study visits.

3. Because of surge in occupancy of hospital beds an extended stay in ER for COVID-19 treatment or home oxygen will qualify for hospital equivalents.

**Medical monitoring**

1. Added independent medical monitor for safety review.

2. Added independent three physician panel to adjudicate COVID-19 related hospitalizations and severity levels of site principal investigators with majority rule decisions.

**Supplement Table 1** Cumulative incidence of grade 3 or 4 adverse events by treatment status

| | **Control (N=589)** | **CCP (N=592)** | **P-Value[1]** |
|---|---|---|---|
| **Number of Grade 3 or 4 events** | **53** | **34** | |
| **Total person-years** | 127.89 | 133.39 | |
| Rate per person-years | 0.41 | 0.25 | |
| Median [IQR} observation time, days | 90 [83,92] | 90 [84,93] | |
| Rate difference (95% CI) | | 0.16 (0.02, 0.30) | 0.03 |

| | **Control (N=589)** | **CCP (N=592)** | **P-value[2]** |
|---|---|---|---|
| **Number randomized, n** | **615** | **610** | |
| **Number transfused, n** | **589** | **592** | |
| **Details of grade 3 or 4 adverse events, n** | | | |
| Pneumonia | 30 (56.6%) | 14 (41.2%) | |
| Bronchial infection | 1 (1.9%) | 0 (0%) | |
| Dyspnea | 1 (1.9%) | 0 (0%) | |
| Hypoxia | 3 (5.7%) | 0 (0%) | |
| Pneumonitis | 2 (3.8%) | 0 (0%) | |

[1] P-value for risk difference between Control and Convalescent
[2] P-values calculated using Fisher's exact test for count data

| | | |
|---|---|---|
| Chest pain - cardiac | 1 (1.9%) | 0 (0%) |
| Hypertension | 1 (1.9%) | 2 (5.9%) |
| Hypotension | 1 (1.9%) | 0 (0%) |
| Non-cardiac chest pain | 1 (1.9%) | 1 (2.9%) |
| Headache | 1 (1.9%) | 0 (0%) |
| Sinus pain | 1 (1.9%) | 0 (0%) |
| Vasovagal reaction | 2 (3.8%) | 4 (11.8%) |
| Infusion related reaction | 1 (1.9%) | 1 (2.9%) |
| Vomiting | 1 (1.9%) | 1 (2.9%) |
| Pancreatitis | 1 (1.9%) | 1 (2.9%) |
| Renal calculi | 1 (1.9%) | 0 (0%) |
| Urinary tract obstruction | 1 (1.9%) | 0 (0%) |
| Leukocytosis | 1 (1.9%) | 0 (0%) |
| Neutrophil count decreased | 1 (1.9%) | 0 (0%) |
| White blood cell decreased | 1 (1.9%) | 0 (0%) |

**Supplement Table 2** Cumulative incidence of severe transfusion reactions by treatment status

| | Control (N=589) | CCP (N=592) | P-Value[3] |
|---|---|---|---|
| **Total severe transfusion reactions** | **0** | **2** | |
| **Total person-years**[4] | 42.03 | 43.32 | |
| Rate per person-years | 0.00 cases/py | 0.05 cases/py | |
| Median [IQR} observation time, days | 28.0 [28.0, 28.0] | 28.0 [28.0, 28.0] | |
| Rate difference (95% CI) | | -0.05 (-0.11, 0.02) | 0.16 |

| | **Control (N=589)** | **CCP (N=592)** | **P-value**[5] |
|---|---|---|---|
| **Number randomized, n** | **615** | **610** | |
| **Number transfused, n** | **589** | **592** | |
| **Number of severe transfusion reactions, n** | **0** | **2** | **0.50** |
| **Details of severe transfusion reactions, n** | | | |
| Pneumonia | 0 (0.0%) | 1 (50.0%) | |
| Infusion related reaction, unspecified | 0 (0.0%) | 1 (50.0%) | |

[3] P-value for risk difference between Control and Convalescent
[4] Participants administratively censored at Day 28
[5] P-values calculated using one-sided Fisher's exact test for count data

**Supplement Table 3** Cumulative incidence of ARDS by treatment status

| | Control (N=589) | CCP (N=592) | P-Value[6] |
|---|---|---|---|
| **Number of ARDS cases** | **1** | **0** | |
| **Total person-years**[7] | 127.89 | 133.39 | |
| Rate per person-years | 0.01 cases/py | 0.00 cases/py | |
| Median [IQR} observation time, days | 90 [83,92] | 90 [84,93] | |
| Rate difference (95% CI) | | 0.01 (-0.01, 0.02) | 0.32 |

[6] P-value for risk difference between Control and CCP
[7] Participants censored at study closeout
